# An empirical study on the effect of PNF training on the recovery of patients with medial collateral ligament injury: Randomized controlled trial

**DOI:** 10.1101/2025.07.05.25330945

**Authors:** Mingming Li, Rui Chu, Xiang Zhang, Yi Peng

**Author notes:** Mingming Li and Rui Chu contributed to the article equally. **Corresponding author**.Xiang Zhang, **Email:** (R.Chu), (X.Zhang). **Fund Project:**2023 discipline (Professional) leader cultivation project “young and middle-aged teachers’ action discipline (Professional) leader cultivation project”(DTR2023071). **Data access links:**10.17632/txfn2hsy7h.l (DOI).

## Abstract

**Objective:** This study aims to investigate the effects of combining medial collateral ligament (MCL) reconstruction surgery with proprioceptive neuromuscular facilitation (PNF) training on knee joint function and rehabilitation outcomes. Based on these findings, relevant clinical recommendations are proposed.

**Methods:** A mixed-method approach was employed, incorporating literature review, experimental design, statistical analysis, and comparative evaluation.

**Results:** Compared to the control group, patients in the PNF group demonstrated significantly greater range of motion (ROM) and knee flexion (P < 0.01), experienced faster reduction in pain (P < 0.01), and showed superior physical recovery (P < 0.01). Overall rehabilitation outcomes were also rated more favorably in the PNF group (P< 0.01).

**Conclusion:** PNF training contributes significantly to the improvement of knee joint stability and motor control in patients with MCL injuries, particularly by strengthening muscles associated with the medial collateral ligament.This clinical trial was registered (https://www.clinicaltrials.gov). The trial number is ChiCTR2500105207.

**Recommendations:** Future research should focus on the effectiveness of PNF training for different types of injuries, explore the impact of varied PNF parameters on MCL rehabilitation, and assess the combined efficacy of PNF with other rehabilitation interventions to maximize its utility in sports medicine.

## 1. Research Background

The medial collateral ligament (MCL) is one of the key stabilizing structures of the knee joint, primarily responsible for resisting valgus stress and external rotational forces ^[1]^. MCL injuries account for approximately 25% to 50% of all knee ligament injuries and are particularly common among athletes involved in high-impact sports such as football and ice skating. These injuries are generally classified into three grades: Grade I (minor sprain or microscopic tears), Grade II (partial tears), and Grade III (complete rupture).

Conservative treatments—such as cold compresses, compression bandages, or immobilization—are commonly prescribed, though the recovery process is often lengthy and prone to complications. In cases of Grade III injuries, surgical reconstruction is typically required; however, postoperative rehabilitation remains a prolonged and complex process. Accelerating recovery and enhancing rehabilitation outcomes is thus a critical concern in clinical practice.

Proprioceptive neuromuscular facilitation (PNF) is a neuromuscular rehabilitation method designed to restore movement function by modulating muscle tone. It improves postural balance and coordination, mitigates the risks associated with poor posture and muscle stiffness, and promotes overall physical recovery ^[2]^. While PNF training has shown considerable efficacy in enhancing motor performance and has been widely adopted in rehabilitation settings ^[3–7]^, few studies have specifically examined its application in patients recovering from MCL injuries.

Therefore, this study seeks to fill that gap by evaluating the effects of PNF training in this patient population, with the goal of improving rehabilitation outcomes and reducing recovery time. The results are expected to provide valuable insights into optimizing treatment strategies and improving patient quality of life.

## 2. Research Subjects and Methods

### 2.1 Subjects

Participants were selected using random sampling from The First Affiliated Hospital of Anhui Medical University; The First Affiliated Hospital of USTC; Wuhu Fifth People’s Hospital (Anhui Wannan rehabilitation hospital) in Anhui Province. The inclusion criteria were as follows:

1. Age between 18 and 45 years;
2. Onset of injury within the past six months, with suboptimal response to prior rehabilitation;
3. Confirmed severe medial collateral ligament (MCL) injury requiring surgical reconstruction.

Exclusion criteria included:

1. History of prior MCL reconstruction;
2. Presence of other severe ligament injuries or fractures in the affected joint;
3. Serious comorbid conditions involving the heart, lungs, liver, or kidneys;
4. Diagnosed cervical spondylosis or other musculoskeletal disorders.

**Figure 1.**
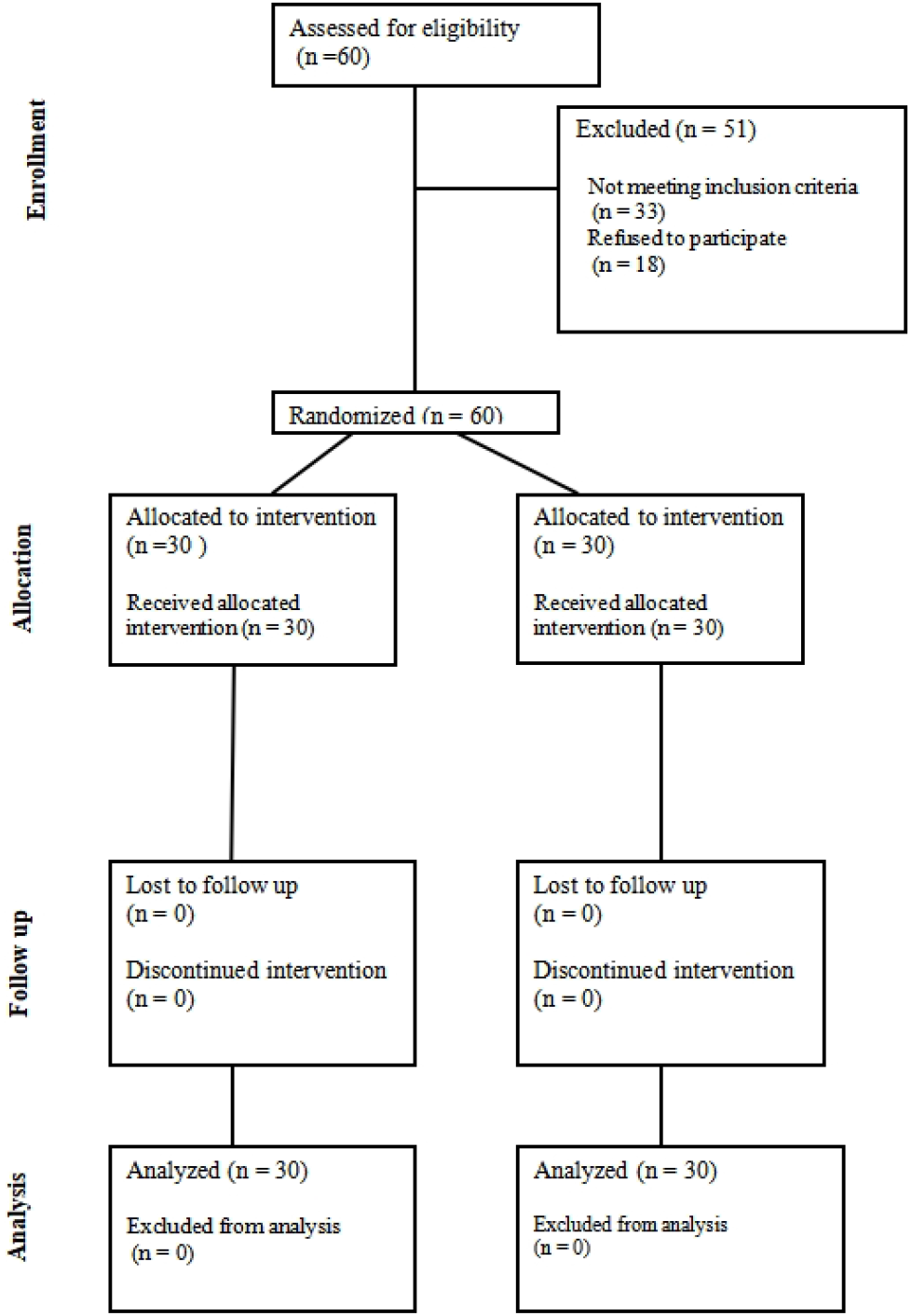
The CONSORT (Consolidated Standards of Reporting Trials) flow chart

### 2.2 Methods

#### 2.2.1 Literature Review

Relevant literature on PNF training in rehabilitation contexts was reviewed using databases such as CNKI, Wanfang Data, and the National Library of China. This review provided a theoretical foundation for the present study.

#### 2.2.2 Experimental Design

1. Research Hypothesis It was hypothesized that integrating PNF training with standard postoperative rehabilitation for MCL reconstruction would yield superior functional outcomes, reduce recovery duration and medical costs, and improve patients’ quality of life. This hypothesis is grounded in the limitations of conventional rehabilitation, which often results in restricted joint mobility and slow early recovery. PNF, by contrast, has demonstrated promising results in neuromuscular rehabilitation.
2. Experimental Parameters and Assessment Measures Key parameters and evaluation indices were selected based on established standards in orthopedic rehabilitation:
  - Demographic and clinical parameters: age, gender, injury severity, surgical method, frequency and duration of PNF sessions, and follow-up intervals.
  - Outcome indicators:
  - Range of Motion (ROM): Assessed by measuring knee flexion and extension before and after surgery.
  - Visual Analog Scale (VAS): A 0–10 scale used to evaluate postoperative pain levels.
  - Lysholm Knee Scoring Scale: A functional assessment tool with a maximum score of 100; higher scores indicate better joint function.
  - Muscle Strength: Assessed using peak torque to determine the degree of functional recovery.
3. Evaluation Methods:
  - ROM: Measured using a goniometer placed between the fibula and tibia on the medial and lateral aspects of the knee.
  - VAS: Pain scores were collected at fixed postoperative intervals using a visual analog scale.
  - Lysholm Score: Functional assessments were based on video recordings and daily rehabilitation logs, evaluating eight dimensions: limp, support, locking, instability, pain, swelling, stair climbing, and squatting.
  - Muscle Strength: Evaluated using isokinetic testing equipment under a controlled speed setting to measure torque output.
4. Experimental Procedure A randomized controlled trial was conducted with 60 patients diagnosed with MCL rupture. Patients were divided into two groups: the experimental group (n = 30) received PNF training following MCL reconstruction, while the control group (n = 30) received standard postoperative rehabilitation without PNF.

PNF Training Protocol:

- Exercise selection: Functional movement patterns within safe limits were chosen to ensure optimal eccentric-to-concentric force transitions.
- Training strategy: PNF methods were selected based on movement nature, intensity, and timing.
- Training frequency: Daily sessions lasting approximately 60 minutes were initiated on the second postoperative day and continued for 12 weeks under the guidance of certified physiotherapists. Emphasis was placed on strengthening the medial musculature, enhancing endurance, and restoring joint stability.

Both groups received equivalent general rehabilitation, including passive movement in the early stage, active movement in the middle stage, and strength training in the later stage. Assessments were conducted at 4, 8, and 12 weeks postoperatively.

#### 2.2.3 Statistical Analysis

Data were analyzed using SPSS 23.0. Paired-sample t-tests were conducted to compare outcome measures between groups. Statistical significance was set at P < 0.01.

#### 2.2.4 Comparative Analysis

Comparative evaluation of outcome measures between the experimental and control groups was used to interpret the effects of PNF training on postoperative recovery.

#### 2.2.5 Ethics approval and consent to participate

The study purpose, method, and procedures were explained to all participants, and they signed a consent form that included information such as the option of voluntary withdrawal from participation.Ethical approval was obtained from the Ethics Committee of Hefei Preschool Teachers College; the approval number is HFYZLZ2025032101. The clinical trial was registered (https://www.clinicaltrials.gov), and the trial number is ChiCTR2500105207.

## 3. Results and Analysis

### 3.1 Descriptive Statistics

A total of 60 patients with medial collateral ligament (MCL) injuries were recruited and randomized into two groups of 30 participants each. The experimental group received standard postoperative care supplemented with PNF training, while the control group received only conventional rehabilitation. All participants underwent MCL reconstruction and were subjected to identical recovery protocols, including early-stage passive movement, mid-stage active movement, and late-stage muscle strengthening. Detailed demographic data and injury characteristics were recorded at baseline. The only difference between groups was the addition of PNF training in the experimental group.

### 3.2 Comparison of Knee Flexion ROM Scores

To evaluate recovery of knee joint range of motion (ROM), knee flexion angles were measured at 4, 8, and 12 weeks postoperatively in both groups. Results are presented in Table 1:

**Table 1.** Comparison of ROM score of knee flexion between the two groups after operation.

| Group | n | Rehabilitation Duration |  |  |
| --- | --- | --- | --- | --- |
|  |  | Week 4 | Week 8 | Week 12 |
| Experimental | 30 | 92.97 ± 4.75 | 122.43 ± 4.53 | 135.20 ± 2.41 |
| Control | 30 | 87.87 ± 3.01 | 106.33 ± 4.95 | 125.03 ± 6.13 |
| t |  | -0.085 | 14.620 | 9.352 |
| Sig |  | 0.003 < 0.01 | 0.000 < 0.01 | 0.000 < 0.01 |

Statistical analysis using paired-sample t-tests revealed that the experimental group had significantly greater knee flexion ROM at all assessment points. The between-group differences increased over time, indicating that PNF training produced a cumulative positive effect on joint mobility.

### 3.3 Comparison of VAS Pain Scores

Pain intensity was evaluated using the Visual Analog Scale (VAS) at 4, 8, and 12 weeks postoperatively. Results are shown in Table 2:

**Table 2.** Comparison of VAS Scores Between Groups.

| Group | n | VAS score (points) |  |  |
| --- | --- | --- | --- | --- |
|  |  | Week 4 | Week 8 | 1Week 12 |
| Experimental | 30 | 1.97±0.98 | 0.79±0.49 | 0.41±0.50 |
| Control | 30 | 3.62±1.74 | 1.65±1.17 | 0.68±0.66 |
| t |  | -4.226 | -3.360 | -1.491 |
| P |  | 0.000 <0.01 | 0.002 <0.01 | 0.001 <0.01 |

Pain levels in the PNF group declined more rapidly and significantly compared to the control group, especially in the early stages of rehabilitation. By week 12, pain had nearly resolved in both groups, but the experimental group exhibited a faster trajectory of relief.

### 3.4 Comparison of Lysholm Functional Scores

The Lysholm Knee Scoring Scale was used to assess knee joint function at 4, 8, and 12 weeks. Results are shown in Table 3:

**Table 3.** Comparison of Lysholm Scores Between Groups.

| Group | n | Muscle strength score (kg) |  |  |
| --- | --- | --- | --- | --- |
|  |  | Week 4 | Week 8 | Week 12 |
| Experimental | 30 | 65.36±2.87 | 80.50±5.07 | 92.33±2.50 |
| Control | 30 | 54.76±9.15 | 70.50±8.80 | 76.76±11.26 |
| t |  | 6.622 | 4.722 | 6.922 |
| P |  | 0.000 <0.01 | 0.000 <0.01 | 0.000 <0.01 |

From week 4 onward, the experimental group consistently demonstrated significantly better functional recovery. By week 12, patients in the PNF group had nearly regained full knee function, while the control group still showed signs of functional limitation.

### 3.5 Comparison of Muscle Strength Scores

Quadriceps and hamstring strength were assessed using isokinetic dynamometry at weeks 4 and 12. Results are presented in Tables 4.

**Table 4.**
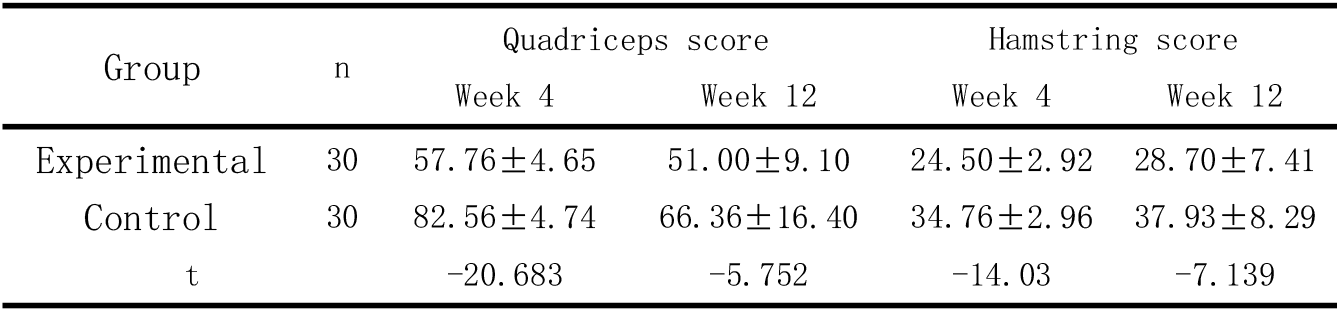
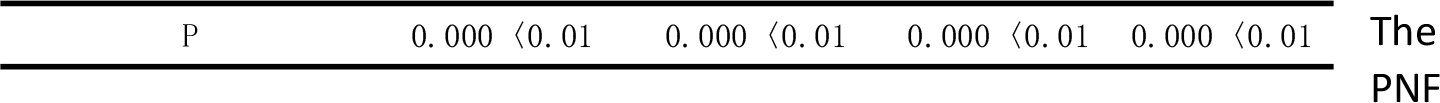
Comparison of Quadriceps and Hamstring Strength Between Groups (kg)

The PNF group showed significantly greater improvements in quadriceps and hamstring strength by week 12. Although the control group also improved, the experimental group achieved closer-to-normal muscle function.

The findings indicate that PNF training significantly enhances postoperative recovery in patients with MCL injuries. Improvements were observed in joint mobility, pain reduction, functional scores, and muscle strength. These results support the clinical use of PNF as a supplementary rehabilitation strategy following MCL reconstruction, corroborating previous research in related domains ^[8–12]^.

## 4. Discussion

This study evaluated the efficacy of proprioceptive neuromuscular facilitation (PNF) training as a postoperative rehabilitation strategy for patients undergoing medial collateral ligament (MCL) reconstruction. While the findings support the clinical value of PNF in enhancing functional recovery, several limitations must be acknowledged, which should be addressed in future research.

### 4.1 Selection of Study Participants

This study exclusively involved patients who had undergone surgical reconstruction of the MCL. Although the results suggest beneficial effects of PNF training in this population, the findings may not generalize to patients with less severe injuries treated conservatively. Further investigation is needed to determine whether PNF training is similarly effective in non-surgical cases or in patients with different grades of MCL injury.

### 4.2 Duration of the Rehabilitation Program

The rehabilitation protocol in this study lasted for 12 weeks. While significant improvements in joint mobility, pain reduction, and muscle strength were observed, long-term outcomes remain unknown. The lack of extended follow-up limits our understanding of whether the observed benefits of PNF are sustained over time.Future studies should incorporate longer rehabilitation periods and post-discharge follow-up to assess the durability of PNF-related improvements.

### 4.3 Assessment Methodology

The current study relied on a single set of validated clinical evaluation tools. While tools such as ROM, VAS, Lysholm score, and isokinetic muscle testing are well established, the inclusion of additional assessment methods—such as gait analysis, electromyography (EMG), or patient-reported quality of life scales—could enhance the robustness and multidimensionality of outcome evaluation. Utilizing a more comprehensive battery of tests would allow for a deeper understanding of the neuromuscular and functional recovery process.

In summary, although the present study demonstrates that PNF training significantly improves recovery following MCL reconstruction, limitations in sample scope, duration, and outcome measurement suggest the need for more comprehensive and longitudinal studies. Expanding the application of PNF beyond surgically treated cases and incorporating multimodal assessment strategies would contribute to a more nuanced understanding of its rehabilitative potential. These findings also offer a meaningful extension to existing rehabilitation literature on MCL injuries, underscoring the importance of evidence-based, individualized recovery protocols ^[13–15]^.

## 5. Conclusion and Recommendations

### 5.1 Conclusion

1. The empirical findings demonstrate that, compared to conventional rehabilitation alone, PNF training significantly improves postoperative outcomes in patients with medial collateral ligament (MCL) injuries. Specifically, patients in the PNF group exhibited greater joint mobility, increased knee flexion angle (P < 0.01), more rapid pain reduction (P < 0.01), and superior gains in muscular strength (P < 0.01). Subjective evaluations of recovery were also more favorable in the PNF group (P < 0.01).
2. PNF training was particularly effective in restoring knee joint stability and enhancing neuromuscular control. Its benefits were most evident in strengthening muscle groups associated with the medial collateral ligament—an outcome that surpasses the effectiveness of traditional rehabilitation modalities alone.

### 5.2 Recommendations

1. Expand research across varying injury types: This study focused on patients undergoing surgical reconstruction for severe MCL injuries. Future studies should explore the effectiveness of PNF training in patients with mild-to-moderate injuries and in those managed with conservative treatment approaches. Such research will allow for a broader assessment of its applicability in diverse clinical scenarios.
2. Investigate parameter-specific training effects: The present study employed a standardized PNF protocol. However, PNF training allows for flexibility in exercise variables such as position, intensity, duration, and rest intervals. Future research should evaluate the impact of different training parameters to optimize PNF program design and maximize rehabilitation outcomes.
3. Explore multimodal rehabilitation strategies: In clinical practice, PNF is often integrated with other therapeutic modalities (e.g., electrotherapy, manual therapy, resistance training). Future trials should assess the synergistic effects of combining PNF with other evidence-based interventions to develop comprehensive and personalized rehabilitation protocols.

## Data Availability

Data access links:10.17632/txfn2hsy7h.l (DOI)

## Acknowledgments

The authors declared no potential conflicts of interest with respect to the research, author-ship, and publication of this article.The research was funded by 2023 discipline (Professional) leader cultivation project “young and middle-aged teachers’ action discipline (Professional) leader cultivation project”(DTR2023071)

## Notes

### Competing Interest Statement

The authors have declared no competing interest.

### Clinical Trial

This clinical trial was registered (https://www.clinicaltrials.gov). The trial number is ChiCTR2500105207.

### Clinical Protocols

https://www.chictr.org.cn/hvshowproject.html?id=279757&v=1.0

### Funding Statement

The author(s) received no specific funding for this work.

### Author Declarations

Ethical approval was obtained from the Ethics Committee of Hefei Preschool Teachers College; the approval number is HFYZLZ2025032101.

